# Changes in pain, mood, and functional connectivity following cryo-auriculotherapy in adults with chronic refractory low back pain: An open-label preliminary study

**DOI:** 10.1101/2024.05.17.24301837

**Authors:** Keith M. Vogt, Abdurrehman Khan, Benedict J Alter, Trent D. Emerick, James W. Ibinson, Ajay D. Wasan, Senthilkumar Sadhasivam, Jacques E. Chelly

## Abstract

**Introduction:** This report describes the effects of auriculotherapy (AT) using cryogenic needles in a small cohort of chronic low back pain patients.

**Methods:** The effects of AT on pain, mood, and functioning were recorded in 10 patients with chronic back pain before and after cryogenic AT using patient-reported outcome instruments: Brief Pain Inventory (BPI), Patient Health Questionnaire (PHQ-9), General Anxiety Disorder (GAD-7), and Patient-Reported Outcomes Measurement Information System (PROMIS-29). In addition, resting-state MRI data collected before and 5-7 days after AT treatment were analyzed for changes in functional connectivity in 4 patients.

**Results:** GAD-7 scores decreased from 10.0 to 7.7 (95% confidence interval of difference= 0.2 – 4.4, p=0.036). Other patient-reported outcomes showed non-significant differences in this pilot cohort. Significant functional MRI connectivity changes were observed in 4 patients, suggesting that AT affected areas of the brain involved in pain processing.

**Conclusion:** Cryogenic AT is a technique that may reduce the psychological burden of low back pain, but further study is needed. Preliminary functional connectivity changes support the concept that the effect of AT is centrally mediated.

## Introduction

Chronic pain is one of our nation’s largest healthcare burdens, financially and societally, and one of the most important causes of opioid use disorder. In the context of the current opioid crisis, considerable research efforts have been focus around developing non-opioid alternative to treat chronic back pain.^1^ Auriculotherapy (AT) is a non-pharmacologic technique.^2^ There is evidence that the use of AT may be used to improve acute pain,^3-5^ xerostomia,^6^ migraine attacks^7^ and anxiety.^8,9^ AT can be based on the use needles,^5^ laser,^10^ electrical stimulation,^3^ acupressure,^11^ and more recently cryogenic needle.^4^ Our team has significant experience with AT, having recently completed two clinical studies using AT, which showed decreased pain and opioid use and improved mood following surgery.^3,5^

Auriculotherapy is an ancient technique initially used to treat back pain and was rediscovered in the 1940s by French physician Dr. Paul Nogier, who postulated that the ear contains a complete representation of the body with the head down.^12^ The theory is that any pathological condition is associated with corresponding changes in the electric properties of specific points of the ear. Auriculotherapy consists of generating action potential at specific locations. These are sent to the brain via carinal nerves. The brain responds by sending impulses to both the ear and the periphery to restore homeostasis.

Several cartographies have been developed, but the most recent and the one adopted by the most acupuncture societies is the one proposed by Dr. Alimi.^13^ It is based on the use of a segmentogram with a corpus collosum as the center of symmetry. Auriculotherapy cartography has been validated by some functional MRI studies Including two focusing on the thumb^14,15^ and one focusing on the knee.^16^ However, the use of FMRI for understanding how global brain connectivity may be modulated by therapeutic AT is understudied. The hypothesis was that the use of AT using cryogenic needles will result in appreciable connectivity changes in brain areas associated with pain processing such as the insula, and the anterior cingulate.

## Methods

This study was reviewed and approved by the University of Pittsburgh IRB (STUDY19030375) before any subject was enrolled. The study was registered with ClinicalTrials.gov (NCT06358287). Patients with refractory chronic low back pain defined as patients with persistent low back pain despite treatment were recruited from the UPMC pain clinic. After signing an informed consent, each patient was scheduled for two resting-state fMRI scans lasting less than 30 minutes, separated by 2-7 days. Cryogenic AT was performed after the first fMRI scan. Prior to each fMRI scan, each patient was asked to complete a Brief Pain Inventory (BPI) Pain Interference,^17^ BPI Severity score,^18^ PROMIS29,^19^ Pain Catastrophizing survey (PCS),^20^ Sleep disturbance,^21^ fatigue,^22^ PHQ0-9,^23^ and GAD-7,^24^ Patient Global Impression of Change (PGIC) questionnaire.^25^ Statistical analyses were performed in SPSS version 28, using p < 0.05 as the threshold for statistical significance, using two-tailed tests.

Auriculotherapy was delivered using a cryo-auriculopunctor, which is a AT technique in which a 2-second focal jet of cold nitrogen gas is used as a cryogenic “needle” to stimulate target points along the auricular cartography. Stimulation is bloodless, colorless, odorless, and causes minimal pain at the site of stimulation. The device used for cryo-AT (shown in Figure1) is a simple design and easy to use.

Based on Alimi D cartography,^13^ the treatment points targeted were pain location, using both mesoderm (Ω2) and L5 spine level (B8); pain pathway at the pontine reticular formation (H13), thalamus (G14), and sensory master point (D17); inter-hemispheric communication at corpus callosum (0,0) and the anterior (I20) and posterior (K4) commissures; pain memory, at the amygdala (F15) and rhinencephalon (C19); and overall inflammation using the ACTH point (I17).

Patients were imaged initially and 2-7 days after AT, using a custom head coil, and an 8-minute blood oxygen-level dependent resting-state functional MRI sequence. Higher-order shimming was applied prior to data acquisition. Functional images had 2 mm isotropic spatial resolution and temporal resolution of 1 s. Gradient-echo field maps were obtained in the same image frame, for subsequent inhomogeneity correction. High-resolution T1-weighted anatomical images were also obtained for subsequent registration. Image analysis was performed in MATLAB using the CONN toolbox version 20b, running on SPM version 12. Image data were preprocessed within CONN, including motion correction and denoising. Seed-based functional connectivity was calculated using a region of interest (ROI) to ROI approach, using a standard brain atlas for ROI definitions and 132 anatomical ROIs were included in the analysis. Group average results were calculated in a paired fashion, with no missing pre- vs. post-data. The group average difference in connectivity was calculated as pre-treatment vs. post-treatment statistical contrasts. Connectivity change was thresholded at the connection level with p < 0.001. In this framework, a positive T-score indicates greater connectivity before treatment (a decrease in connectivity after treatment).

## Results

The study group was composed of 3 male and 7 female. All were Caucasian. Mean age was 49.6 ± 15.1 year, BMI 32.0 ± 7.1. All patient reported outcome scores were normally distributed, and are reported as pre- vs. post-treatment average values across patients. BPI pain severity scores were 5.7 vs. 4.8 (p=0.070). BPI pain interference scores were 6.2 vs. 5.4 (p = 0.164). PHQ-9 scores were 13.9 vs. 12.9 (p=0.266). GAD-7 scores significantly improved, from 10 to 7.7 (p=0.036, 95% confidence limits of difference 0.193 to 4.41). Patient global impression change scores were 4.7 vs. 3.9 (p=0.121)

PROMIS-29 anxiety scores were 10.4 vs. 9.6 (p=0.553). PROMIS-29 depression scores were 11.2 vs. 10 (p=0.193). PROMIS-29 sleep disturbance scores were 13.7 vs. 13.2 (p=0.343). PROMIS-29 social functioning scores were 8 vs. 9.2 (p=0.373). PROMIS-29 pain interference scores were 15 vs. 14.8 (p=0.838). PROMIS-29 pain intensity scores were 6.2 vs. 5.6 (p=0.193). Finally, PROMIS-29 pain catastrophizing scores were 27.7 vs. 26.7 (p=0.598).

A preliminary analysis of functional connectivity was performed, based on 4 patients imaged before and 5 days after receiving cryo-AT. Table 1 lists the connections with significant changes. Figure 2 shows a connectome ring displaying significant changes in brain connectivity, comparing the pre-AT to post-AT scans. The figure inlay shows the anatomical location of the center of the ROI on a semi-transparent brain image. These results should be considered hypothesis-generating, as these differences are based on a small sample size and are not robustly corrected for multiple comparisons.

**Table 1:** Changes in connectivity, listing Regions of Interest (ROI) between which connectivity changes were detected, comparing before vs. after auriculotherapy treatment. Abbr= abbreviated name of ROI as shown in Fig. 2.

| ROI 1 |  | ROI 2 |  | Connectivity change Stats |  |
| --- | --- | --- | --- | --- | --- |
| Abbr | Full Name | Abbr | Full Name | T-score | p-unc |
| Cuneal l | Cuneal Cortex Left | OP l | Occipital Pole Left | 45.0 | 0.00002 |
| PaCiG r | Paracingulate Gyrus Right | MidFG r | Middle Frontal Gyrus Right | 20.4 | 0.00026 |
| IFG tri r | Inferior Frontal Gyrus, pars triangularis Right | Ver9 | Cerebellar Vermis 9 | -19.9 | 0.00028 |
| aSMG r | Supramarginal Gyrus, anterior division Right | IFG oper r | Inferior Frontal Gyrus, pars opercularis Right | 19.0 | 0.00032 |
| CO r | Central Opercular Cortex Right | Ver45 | Cerebellar Vermis 4 & 5 | 17.3 | 0.00042 |
| Accumbens r | Nucleus Accumbens Right | Cereb10 l | Cerebellum 10 Left | -16.0 | 0.00053 |
| pTFusCl | Temporal Fusiform Cortex, posterior division Left | Cereb10 r | Cerebellum 10 Right | -15.1 | 0.00064 |
| FP r | Frontal Pole Right | Ver9 | Cerebellar Vermis 9 | -14.5 | 0.00071 |
| Hippocampus l | Hippocampus Left | FP l | Frontal Pole Left | 13.0 | 0.00098 |

**Figure 1:**
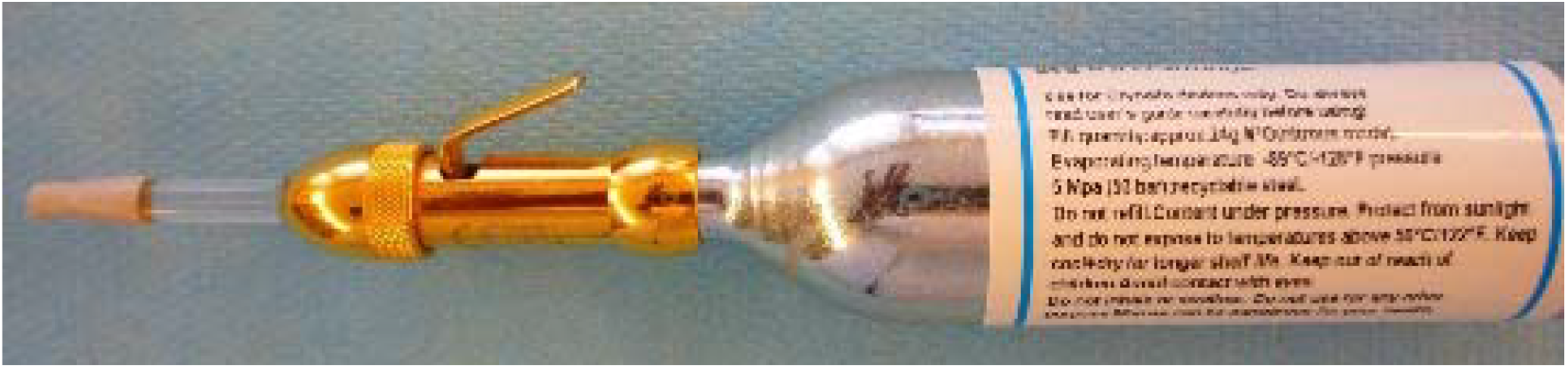
Cryo-auriculopunctor, with compressed nitrogen cannister attached.

**Figure 2.**
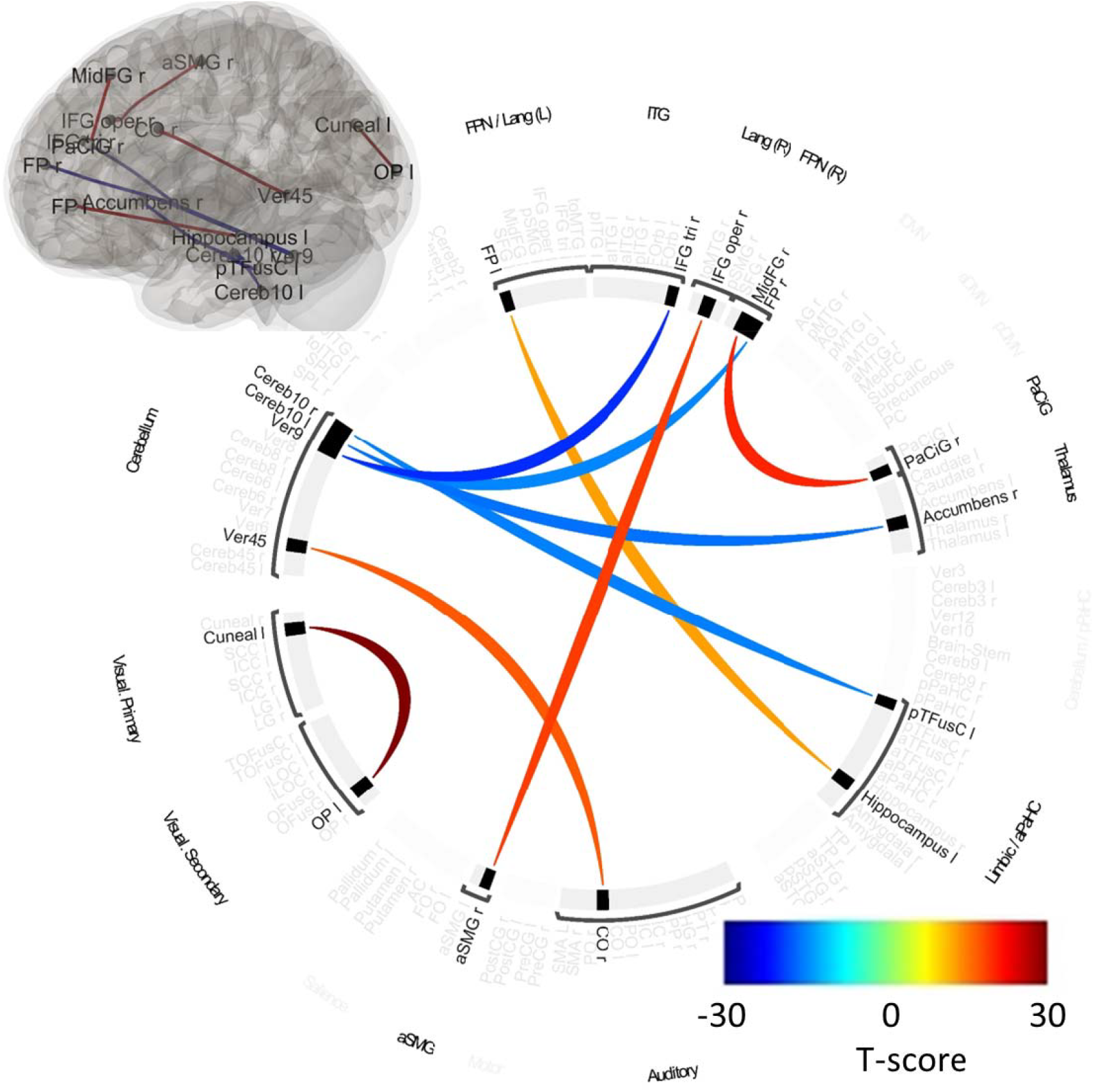
presents the changes in functional connectivity shown as a connectome ring, with color-bar indicating T-score for the connectivity change contrast. The inlay in upper left shows the location of the center of each region on the brain anatomy.

## Discussion

Based on BPI Pain interference and BPI Pain Severity scores recorded before AT, the studied patients suffered from moderate to severe pain. Their level of anxiety and depression was moderate as was their level of fatigue. Catastrophizing scores were borderline normal. In this patient population AT using cryogenic needles appeared to be effective to reduce anxiety when it was assessed GAD-7. However, the magnitude of the changes reported were overall small and variable, consistent with differences expected in a preliminary cohort. Another possible explanation is that PROMIS assessment of pain, anxiety and depression is based on fewer questions than PHQ-9 and GAD-7 reducing granularity of assessment.

Points of cryo-AT were chosen based on the use of Alimi’s cartography. This cartography was validated by the World Federation of Chinese Medicine Societies (WFCMS) during its 8th Convention in London, 2011, which include the participation of Dr. Qi Zhang, MD, who was coordinating of the Traditional Medicine Department, at the World Heath Organization (WHO).^13^ Many other ear cartographies have been proposed by different authors including Nogier P (1966, 1990), Romoli M (1989 and 2009), Orleson T.(1983, 1996), and Wojak W (2004), by different acupuncture societies like the Chinese or European acupuncture society. The premises to the development of these cartographies is based on the use of points or zone, in some cases the use of model allowing to normalize the position of each point such as the segmentogram (Romoli M and Alimi D) and the use of specific center of symmetry such as the point zero (Romoli M) in reference to acupuncture or the corpus collosum (Alimi D) in reference to the center of symmetry of the brain. However, to date none of these cartographies have been validated by a similar large number of representatives from acupuncture societies or by the use of fMRI. In fact a recent comparison of the ear point of the knee using Alimi vs a Chinese/German cartography demonstrated that Alimi cartography was more accurate.^15^

Our data suggest that using Dr Alimi’s cartography and an approach based on the pathophysiology of low back pain, AT may represent an alternative to not only control pain associated with back pain but also mood disorders often associated with this condition. Also in this study, it is important to recognize that AT was only used once. Practically, the use of AT for the treatment of chronic conditions including pain required multiple AT sessions. It is considered that at least 6 treatments will allow to determine the maximum results that can be expected from the use of AT. These treatments are initially spaced 4 weeks apart and after few treatments the patient may experience a longer pain-free interval and therefore the practitioner may offer a longer period of time between treatment. Ultimately, some patients may observe their pain to be very well controlled without any additional treatment after 6 months to 1 year.

Our preliminary results, though from a very small cohort, showed significant changes in fMRI-based functional connectivity after cryo-AT treatment. Connectivity changes were seen between the prefrontal cortex and important areas known to be involved in pain-processing (anterior cingulate) and memory (hippocampus). Interestingly, there were several changes in connectivity detected that involved the cerebellum. The role of the cerebellum in many cognitive functions, including the processing of pain is emerging, but still incompletely understood. However, these findings should be viewed as hypothesis-generating, not definitive. Further research is needed to determine if these preliminary changes persist in a larger study cohort. This also demonstrates the potential role fMRI can play in tracking brain changes over time, in response to treatments for chronic pain.

## Data Availability

All data produced in the present study are available upon reasonable request to the authors.

